# Trends in SARS-CoV-2 detection during social relaxation measures over ten months of COVID-19 pandemic in the metropolitan area of Rio de Janeiro, Brazil

**DOI:** 10.1101/2021.03.02.21252759

**Authors:** Fábio de O. M. Alonso, Bruno D. Sabino, Marcia Soraya C. de Oliveira, Fabiana B. Knackfuss, Rafael B. Varella

**Author notes:** **Address for correspondence:** Dr. Rafael Brandão Varella. Laboratory of Virology, Biomedical Institute-UFF, Brazil. 24210-130.

## Abstract

We analyzed the effects of sequential reopening events during COVID-19 pandemic, based on 76,419 SARS-CoV-2 molecular tests performed from April 2020 to January 2021 in Rio de Janeiro metropolitan area, Brazil, third largest in South America. Post-opening events provoked different impacts on cases and deaths, but showing limited temporary effect.

---

Several authors have assessed the impact of social relaxation events in the COVID-19 pandemic before vaccination was implemented. In general, the proposed analyzes are based on projections by epidemiological models (*1,2*), shorter follow-up periods (*3*), or using data from government databases (*4*). Such information is crucial, providing projections on the pandemic in different scenarios. However, model projections can be conflicting (*5*) or do not correspond to the observed reality (*6*), and short assessments based on secondary data can generate biased analyzes. On the other hand, empirical data obtained from a primary source can provide a closer approximation to reality (*7*), although they are time consuming and carried out *a posteriori*.

Brazil was one of the most affected countries by COVID-19 pandemic (*8*). Its enormous territory combined with variable social conditions, and the adoption of decentralized policies, produced a complex epidemiological scenario (*9*). Rio de Janeiro is the second most populated metropolitan area in Brazil, third in South America, characterized by its high population density, huge variation in income and access to health services. The area accumulated about 348,000 cases of SARS-CoV-2 in period studied (*10*). In mid-March 2020, the state adopted contingency measures that intensified as the pandemic unfolded, gradually reopening circulation from May 5 (epidemiological week, EW, 20) onward. This study allowed us to identify which events did or did not impact the series of SARS-CoV-2 and their temporality.

## The study

We assess trends in cases of SARS-CoV-2 considering the opening events (Appendix) implemented over ten months of COVID-19 pandemic [April/2020 (EW17) to January/2021 (EW4)]. To this end, we rely on the temporal analysis of 76,419 molecular tests for SARS-CoV-2 (Appendix) applied in residents and workers of the metropolitan area of Rio de Janeiro.

Statistical analyzes included Rt data over 41 weeks (EW17/2020 - EW4/2021), considering the number of positive, negative and total cases in the database. The confirmed cases in the laboratory per EW were listed and plotted. For comparison purposes, the official data for the metropolitan area were plotted considering cases and deaths. Descriptive statistical analyzes were performed using SPSS 25.0 (IBM). Initially, the frequency of positive and negative cases was compared by EW using the chi-square test for bilateral significance test with a significance level of 0.01. Subsequently, the chi-square test for trend was used to verify fluctuations in frequencies over a set of weeks.

The percentile value of the number of positives varied from 4.1% (EW41) to 59.3% (EW17) and the highest number of positives were EW19, 18 and 48, respectively. Also, the temporal evolution of new cases varied between EW (Table 1). It was possible to observe an increase with a significant trend (p <0.01) of cases (EW20-22, EW 29-31, EW 41-43, EW 44-46) and a decrease (EW 38-40, EW 50-52, EW 53-4/2021), indicating that the opening events resulted in temporary and non-cumulative effects (Table 2 and Figure). Opening events that occurred in EW20, EW30, EW41 and EW45, perhaps combined with other factors, may have contributed for the upward trend in the set of weeks following these events.

**Table 1.** SARS-CoV-2 cases detected by epidemiological week in metropolitan area of Rio de Janeiro-Brazil

| Epidemiological week | Not detected | Detected | Total | % total | Chi Square | P value |
| --- | --- | --- | --- | --- | --- | --- |
| 17 | 127 | 185 | 312 | 59.3 | 55 | <0.01 |
| 18 | 674 | 514 | 1188 | 43.3 | 49.0954 | <0.01 |
| 19 | 1229 | 769 | 1998 | 38.5 | 527.4324 | <0.01 |
| **20** | 1570 | 393 | 1963 | 20.0 | 74.9224 | <0.01 |
| 21 | 1191 | 413 | 1604 | 25.7 | 33.8257 | <0.01 |
| 22 | 1018 | 365 | 1383 | 26.4 | 775.3159 | <0.01 |
| **23** | 2179 | 227 | 2406 | 9.4 | 376.0861 | <0.01 |
| 24 | 1515 | 152 | 1667 | 9.1 | 286.0496 | <0.01 |
| 25 | 1397 | 131 | 1528 | 8.6 | 214.9698 | <0.01 |
| 26 | 1557 | 168 | 1725 | 9.7 | 236.6959 | <0.01 |
| 27 | 2254 | 245 | 2499 | 9.8 | 262.5591 | <0.01 |
| 28 | 2679 | 251 | 2930 | 8.6 | 140.5692 | <0.01 |
| 29 | 2137 | 218 | 2355 | 9.3 | 182.9477 | <0.01 |
| **30** | 2165 | 167 | 2332 | 7.2 | 56.4049 | <0.01 |
| 31 | 1544 | 169 | 1713 | 9.9 | 29.2377 | <0.01 |
| 32 | 1221 | 147 | 1368 | 10.7 | 20.2133 | <0.01 |
| 33 | 1364 | 179 | 1543 | 11.6 | 14.2454 | 0.0002 |
| **34** | 1352 | 185 | 1537 | 12.0 | 11.1775 | 0.0008 |
| 35 | 1597 | 228 | 1825 | 12.5 | 46.6116 | <0.01 |
| 36 | 1976 | 219 | 2195 | 10.0 | 24.0065 | <0.01 |
| 37 | 1699 | 207 | 1906 | 10.9 | 41.6312 | <0.01 |
| 38 | 1831 | 194 | 2025 | 9.6 | 63.7629 | <0.01 |
| 39 | 1567 | 129 | 1696 | 7.6 | 120.1396 | <0.01 |
| 40 | 1524 | 73 | 1597 | 4.6 | 117.5575 | <0.01 |
| **41** | 1464 | 63 | 1527 | 4.1 | 38.5327 | <0.01 |
| 42 | 1351 | 114 | 1465 | 7.8 | 93.9699 | <0.01 |
| 43 | 1701 | 95 | 1796 | 5.3 | 68.5363 | <0.01 |
| 44 | 1525 | 94 | 1619 | 5.8 | 21.1144 | <0.01 |
| **45** | 1348 | 126 | 1474 | 8.5 | 1.805 | 0.18 |
| 46 | 1386 | 220 | 1606 | 13.7 | 81.5763 | <0.01 |
| 47 | 2023 | 470 | 2493 | 18.9 | 45.0609 | <0.01 |
| 48 | 2476 | 512 | 2988 | 17.1 | 40.7328 | <0.01 |
| 49 | 2137 | 451 | 2588 | 17.4 | 110.0876 | <0.01 |
| 50 | 1745 | 463 | 2208 | 21.0 | 26.9376 | <0.01 |
| 51 | 2088 | 431 | 2519 | 17.1 | 0.0087 | 0.92 |
| 52 | 2076 | 327 | 2403 | 13.6 | 1.2138 | 0.27 |
| 53 | 1201 | 174 | 1375 | 12.7 | 2.4302 | 0.12 |
| 1 | 1637 | 232 | 1869 | 12.4 | 20.9581 | <0.01 |
| 2 | 1902 | 216 | 2118 | 10.2 | 72.219 | <0.01 |
| **3** | 1701 | 122 | 1823 | 6.7 | 58.2821 | <0.01 |
| 4 | 1178 | 75 | 1253 | 6.0 |  |  |
| TOTAL | 66306 | 10113 | 76419 | - | - | - |

\*the epidemiological weeks in bold correspond to the opening dates

**Table 2.** Trend analysis of SARS-CoV-2 detection per epidemiological week in the metropolitan area of Rio de Janeiro-Brazil

| Epidemiological week | A | Trend | Chi Square | p |
| --- | --- | --- | --- | --- |
| **20**, 21, 22 | 88,31 | A>0<br>(growing) | 12,47 | 0,0004 |
| **23**, 24, 25 | -14,7 | A<0<br>(decreasing) | 0,68 | 0,41 |
| 26, 27, 28 | -26,39 | A<0<br>(decreasing) | 36,2 | 0,17 |
| 29, **30**, 31 | 124,2 | A>0<br>(growing) | 36,9 | <0,0001 |
| 32, 33, **34** | 16,35 | A>0<br>(growing) | 0,89 | 0,34 |
| 35, 36, 37 | -26,96 | A<0<br>(decreasing) | 27,81 | 0,16 |
| 38, 39, 40 | -88,95 | A<0<br>(decreasing) | 27,81 | <0,0001 |
| **41**, 42, 43 | 51,27 | A>0<br>(growing) | 15,13 | <0,0001 |
| 44, **45**, 46 | 116,32 | A>0<br>(growing) | 48,87 | <0,0001 |
| 47, 48, 49 | -30,46 | A<0<br>(decreasing) | 1,21 | 0,87 |
| 50, 51, 52 | -144,63 | A<0<br>(decreasing) | 31,03 | <0,0001 |
| 53 ,1, 2, **3**, 4 | -255,09 | A<0<br>(decreasing) | 53,00 | <0,0001 |

\*the epidemiological weeks in bold correspond to the opening dates.

**Figure.**
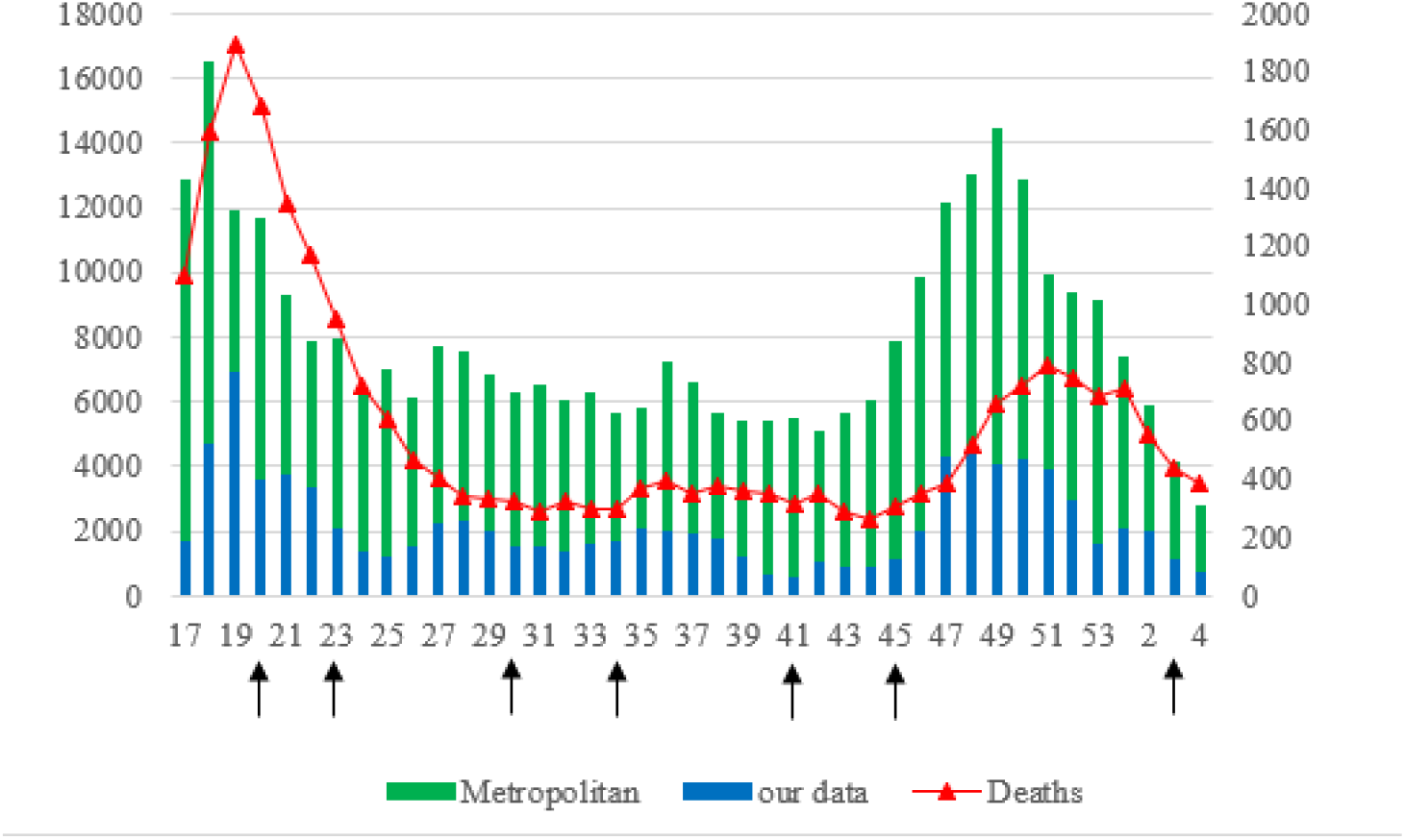
Temporal trend of confirmed cases and deaths by COVID-19 in the metropolitan region of Rio de Janeiro.

^*^Arrows correspond to opening epidemiological weeks

Comparing the official government data of COVID-19 cases over the EW, it was possible to observe that the data from this study responded more accurately to the reopening events (Figure). Official deaths followed the trend of cases, with two prominent peaks in April/May and November/December, which correspond to the beginning of the pandemic and a resurgence in the cases, respectively. Between these two peaks (May to October), several relaxation measures were taken, although deaths remain stable (median of 473; IQR: 454-515).

## Conclusions

In contrast to different authors (*1,3,4,11*), our data showed that most social relaxing events including the opening of bars and restaurants, gyms, beauty salons, shopping centers, transportation, among others (EW 20 to 41), had a limited effect on SARS-CoV-2 cases. Although it was possible to observe an increase in cases after some of the opening events, our results indicate that such measures had a temporary and not a cumulative effect, which was also reflected in the stability of reported deaths, the most important and stable outcome, not subject to testing variations, testing policy and infrastructure (*8*). We also corroborate the results obtained by a modeling study that indicated a delay of 2 to 3 weeks in the increase of cases after opening (*1*), following a drop of 3-4 weeks after the peak, which was confirmed by the trend analysis (Table 2).

After November (EW 45), however, we observed a significant increase in cases (a “second wave”), also seen across the country. Although we cannot pinpoint the specific cause, the release of festive and social events of high agglomeration in closed places such as cinema and theater rooms, parties, birthdays, concerts, etc., in concomitance with the national electoral campaign for mayors and councilors, may have been crucial to this phenomenon. In addition, the increased flexibility of the quarantine in several states and municipalities in Brazil, favored physical isolation rates returned to pre-pandemic levels (*12*).

Not least, the emergence of SARS-CoV-2 variants were detected in the state at the time (*13*), although its transmissibility has not been proven to be major, nor has it been a homogeneous event in the country. It is possible that the sum of all these events was responsible for the “dark November”. Even in this most dramatic scenario, after about 3 weeks, cases started to fall as previously, even without the imposition of a lockdown. The reasons for this temporal pattern are not entirely clear but are probably due to characteristics of the transmission dynamics of the virus, network of social contacts, environment, immunity, and others (*14*). We hypothesized the first opening events kept the social network relatively stable within the communities *(15)*, preventing a significant increase in cases. This situation was disturbed by the post-EW 45 events, where an overlap of social groups occurred, leading to an explosion of cases. Nonetheless, this temporary post-opening pattern is difficult to observe from projection analysis (*6*).

A closer trend was observed between our data and the relaxation measures implemented. This higher sensitivity was probably due to the use of molecular methodology throughout the research, while official data is based on different diagnostic approaches, including rapid tests and clinical examination, less accurate than PCR-based tests.

Our study has limitations. We did not directly observe the effects of social relaxation on hospitalizations and their outcomes. As previously stated, variations in testing availability, groups and geographical areas, also cause artificial variations in frequency of viral detection. The opening of schools, considered a relevant aspect in the pandemic according to some models (*1*), was not evaluated because it remained closed during the study. In addition, municipalities had some flexibility in decision-making, not always strictly following state decrees.

While vaccination against SARS-CoV-2 is unfolding worldwide, doubts on social relaxation strategies remain due to conflicting or scarce data. In fact, it is still unclear which events significantly impact the pandemic. To the best of our knowledge, this is one of the longest follow-up of reopening events during the COVID-19 pandemic based on empirical data. Our study provides valuable data on the case series of SARS-CoV-2 after a sequence of social relaxation events, which can be useful for decision making.

## Data Availability

The authors confirm that the data supporting the findings of this study are available within the article and its supplementary materials

## Acknowledgments

Varella RB was partially supported by National Council for Scientific and Technological Development – CNPq. The authors report no conflicts of interest.

## Notes

### Competing Interest Statement

The authors have declared no competing interest.

### Author Declarations

The study was approved by the University Hospital Ethical Committee of the Fluminense Federal University (register 30926020.2.0000.5243)

## References

1. Li Y, Campbell H, Kulkarni D, Harpur A,Nundy M,Wang X, et al. The temporal association of introducing and lifting non-pharmaceutical interventions with the time-varying reproduction number (R) of SARS-CoV-2: a modelling study across 131 countries The Lancet.Infect Dis. 2020 Oct;21(2):193–202. https://doi.org/10.1016/S1473-3099(20)30785-4

2. Kissler SM, Tedijanto C, Goldstein E, Grad YH, Lipsitch M. Projecting the transmission dynamics of SARS-CoV-2 through the postpandemic period. Science. 2020 May 22;368(6493):860–868. https://doi: 10.1126/science.abb5793

3. Tsai AC, Harling G, Reynolds Z, Gilbert RF, Siedner MJ. COVID-19 transmission in the U.S. before vs. after relaxation of statewide social distancing measures. Clin Infect Dis. 2020 Oct 3:ciaa1502. https://doi: 10.1093/cid/ciaa1502

4. Kaufman BG, Whitaker R, Mahendraratnam N, Smith VA, McClellan MB. Comparing Associations of State Reopening Strategies with COVID-19 Burden. J Gen Intern Med. 2020 Dec;35(12):3627–3634. https://doi: 10.1007/s11606-020-06277-0

5. Rice K, Wynne B, Martin V, Ackland GJ. Effect of school closures on mortality from coronavirus disease 2019: old and new predictions. BMJ. 2020 Oct 7;371:m3588. https://doi: 10.1136/bmj.m3588

6. Ioannidis JPA, Cripps S, Tanner MA. Forecasting for COVID-19 has failed. Int J Forecast. 2020 Aug 25. https://doi: 10.1016/j.ijforecast.2020.08.004

7. Campello de Souza, Bruno and Campello de Souza, Fernando Menezes, Does Social Isolation Really Curb COVID-19 Deaths? Direct Evidence from Brazil that it Might do the Exact Opposite. http://dx.doi.org/10.2139/ssrn.3706464

8. World Health Organization. Coronavirus disease (COVID-19) dashboard. [cited 2020 Nov. 20]. https://covid19.who.int/table

9. Cavalcante JR, Cardoso-Dos-Santos AC, Bremm JM, Lobo AP, Macário EM, Oliveira WK, França GVA. COVID-19 in Brazil: evolution of the epidemic up until epidemiological week 20 of 2020. EpidemiolServSaude. 2020;29(4):e2020376. http://10.5123/s1679-49742020000400010

10. CoronavirusPanel RJ. Rio de Janeiro state, Brazil. [cited 2021 Feb 20]. http://painel.saude.rj.gov.br/monitoramento/covid19.html#

11. Hussein NR, Naqid IA, Saleem ZSM, Almizori LA, Musa DH, Ibrahim N. A sharp increase in the number of COVID-19 cases and case fatality rates after lifting the lockdown in Kurdistan region of Iraq. Ann Med Surg (Lond). 2020 Jul 24;57:140–142. https://doi: 10.1016/j.amsu.2020.07.030

12. AsaiG, Kuroiva A, Terra ML. Brazilian model estimation for SARS-CoV-2 peak contagion (BMESPC). Medrxiv. 2021 (6): 1–18. https://doi.org/10.1101/2021.01.02.20248940

13. Voloch C, Ronaldo F, Almeida L, Cardoso C, Brustolini O, Gerber A, et al. Genomic characterization of a novel SARS-CoV-2 lineage from Rio de Janeiro, Brazil. Medrxiv. 2020. https://10.1101/2020.12.23.20248598

14. Thurner S, Klimek P, Hanel R. A network-based explanation of why most COVID-19 infection curves are linear. Proc Natl Acad Sci U S A. 2020 Sep 15;117(37):22684–22689. https://doi: 10.1073/pnas.2010398117

15. Li Y, Wang X, Nair H. Global Seasonality of Human Seasonal Coronaviruses: A Clue for Postpandemic Circulating Season of Severe Acute Respiratory Syndrome Coronavirus 2? J Infect Dis. 2020 Sep 1;222(7):1090–1097. https://doi: 10.1093/infdis/jiaa436

