## Appendix for "Trends in SARS-CoV-2 detection during social relaxation measures over ten months of COVID-19 pandemic in the metropolitan area of Rio de Janeiro, Brazil"

**Methods**

1) Reopening measures in Rio de Janeiro state during COVID-19 pandemic.

After the peak, in the April-May transition (EW 18), social relaxation events were gradually and cumulatively implemented from May 2020 (EW 20), based on varied epidemiological criteria, which served as a basis for the reopening of the municipalities. Until the closing of the data for this article (EW 4 of 2021), schools were not yet fully functioning in the state and vaccination campaigns were yet to begin. Mandatory use of masks in public and closed places remained unchanged for the entire period studied. The main social relaxation events were roughly decreed on a monthly basis, and their concentration period are summarized in Supplementary Table XX.

Official data of COVID-19 cases and deaths in Rio de Janeiro state is available at: http://painel.saude.rj.gov.br/monitoramento/covid19.html#

2) Laboratorial analyses

Samples were extracted by Quick-DNA/RNA™ Viral MagBead automated kit (Zymo Research, CA, USA) and tested for SARS-CoV-2 by RT-qPCR using Allplex™2019-nCoV Assay (Seegene Inc., Seoul, Korea). The tested population was heterogeneous, being composed of individuals suspected of COVID-19, close contacts, as well as those submitted to screening for work activities.

Supplementary Table 1. Main reopening events according to epidemiological week.

| EW | reopening characteristics |
|---|---|
| 20 (11/05) | operation of small establishments, markets and supermarkets allowed, being prohibited the continued permanence and the agglomeration of people in these places. |
| 23 (05/06) | Intercity transportation allowed with 50% of its capacity; Bars, restaurants and shopping centers, with a limit of 50% of its capacity and time limitation; Few touristic spots opened; Religious organizations can work, as long as the distance of 1 meter between people is observed; Outdoor activities allowed as long as there is no agglomeration; Cultural activities of any nature in the drive-in model; Free fairs for food products, with restrictions such as the distance of stalls of 1 meter; Availability of 70% alcohol in all the aforementioned locations and events. (1st opening entertainment, transport, religious activities) |
| 30 (22/07) | Reopening of new sectors of commerce, such as gyms and beauty salons, if protocols and security measures are respected; Outdoor cultural activities in the metropolitan, coastal and northwest areas, |

| | |
|---|---|
| | respecting the minimum distance of one meter between people; reopening of more public agencies. |
| 34 (19/08) | Reopening of cultural establishments in the regions where the risk of contamination is considered low; social events in environments such as ballrooms and party houses, as long as the access to the interior of the establishment is restricted, respecting the limit of 1/3 of the total capacity limit of the place; partial resumption with 1/3 (one third) of occupations of theater rooms, concert halls, museums and cultural centers in the state of Rio de Janeiro; holding business, corporate and scientific events in environments such as hotels, pavilions, convention center and the like, provided that access contention is ensured inside the establishment, respecting the limit of 1/3 (one-third) of the total site capacity limit |
| 41 (06/10) | partial resumption of the activities of the Itinerant Amusement Parks, as long as they are respected strictly comply with the rules of social distancing, using 50%; events should take place in open outdoor spaces whose total capacity for public is equal to or greater than 3 thousand people (three thousand people), respecting the maximum limit of<br>1/3 (one third) of the capacity of the site. |
| 45 (05/11) | Release for social events in general (cinema and theater rooms, parties, concerts, events, shows, etc.), with about 50% or 1/3 of the capacity; Social events, such as weddings, weddings, birthdays, graduations, cocktails,gatherings, inaugurations, launches, official ceremonies, among others that follow this same format, will be allowed with the limitation of 50% of the audience capacity; Samba School courts and Carnival Blocks headquarters may hold events |
| 03 (21/01) | Maintenance of the EW45 measures, with expansion of the public to 1/2 or 2/3 of the capacity of the site. |

[1]Although municipalities had decision autonomy, the metropolitan area RJ followed these criteria with little temporal variation.EW: epidemiological week.

Supplementary Table 2. COVID-19 cases and deaths by official data in the metropolitan area of Rio de Janeiro-Brazil.

| Epidemiological week | Cases | Deaths |
|---|---|---|
| 17 | 2285 | 1142 |
| 18 | 3718 | 1658 |
| 19 | 6383 | 1986 |
| 20 | 4672 | 1806 |
| 21 | 12932 | 1473 |
| 22 | 17887 | 1315 |
| 23 | 12113 | 1120 |
| 24 | 14303 | 865 |
| 25 | 16701 | 749 |
| 26 | 13266 | 625 |
| 27 | 11637 | 579 |

| | | |
|---|---|---|
| 28 | 9235 | 515 |
| 29 | 5555 | 525 |
| 30 | 21063 | 515 |
| 31 | 10920 | 457 |
| 32 | 11311 | 473 |
| 33 | 12090 | 443 |
| 34 | 19850 | 454 |
| 35 | 12493 | 509 |
| 36 | 9790 | 520 |
| 37 | 8029 | 462 |
| 38 | 10485 | 496 |
| 39 | 8409 | 456 |
| 40 | 11905 | 438 |
| 41 | 11832 | 399 |
| 42 | 6162 | 430 |
| 43 | 9254 | 356 |
| 44 | 11154 | 345 |
| 45 | 5371 | 389 |
| 46 | 11297 | 445 |
| 47 | 10632 | 483 |
| 48 | 15483 | 645 |
| 49 | 17507 | 820 |
| 50 | 18164 | 905 |
| 51 | 15229 | 992 |
| 52 | 16415 | 977 |
| 53 | 15529 | 919 |
| 1 | 24262 | 918 |
| 2 | 21245 | 762 |
| 3 | 19074 | 623 |
| 4 | 20545 | 530 |
